# Diet Quality Scores are Associated with Sleep Timing and Regularity in Emerging Adults

**DOI:** 10.64898/2026.10.25.26366435

**Authors:** Sofia O. Sanchez, Meral N. Culver, Braxton A. Linder, Zach J. Hutchison, Sarah M. Honaker, Nathaniel D.M. Jenkins, Ellen M. Evans, Austin T. Robinson

## Abstract

**Background:** Sleep health is associated with cardiometabolic health, potentially due to direct physiological effects, altered diet quality, or a combination of both. However, data on multidimensional sleep health, particularly sleep regularity and holistic dietary patterns, in emerging adults remain limited.

**Objective:** To examine associations between sleep health and diet quality in emerging adults.

**Methods:** Objective sleep (duration, efficiency, timing [bedtime, waketime, midpoint], regularity) was measured via actigraphy: n=106 (48M/58F, age [median = 22, IQR = 2] years, BMI [24.2, 4] kg/m^2^). Subjective sleep quality was measured via Pittsburgh Sleep Quality Index: n=122 (58M/64F, age [22, 2], BMI [24.2, 5] kg/m^2^. We collected diet data via daily food and fluid diaries (≥3 days) to determine Dietary Approaches to Stop Hypertension (**DASH**; 0-9) and Healthy Eating Index (**HEI**; 0-100) scores. We used hierarchical regressions (model 1: bivariate analysis, model 2: sex, BMI, race) to examine associations between sleep health and diet quality. We compared diet quality between participants obtaining adequate and inadequate sleep for each sleep health metric using Mann-Whitney U tests.

**Results:** Sleep midpoint and bedtime were independently associated with DASH (Bs ≥ -0.165, ps ≤ 0.001) and HEI (Bs ≥ -2.180, ps ≤ 0.001) after inclusion of covariates. Group comparisons indicated higher diet quality among participants with adequate sleep midpoint and waketime SD (*p* ≤ 0.040).

**Conclusions:** Higher sleep regularity is associated with better diet quality in emerging adults. These relations may be bidirectional, thus future studies are needed to investigate whether sleep health interventions improve diet quality, or vice versa, among emerging adults.

## Introduction

Sleep health is increasingly recognized for its impact on cardiometabolic health (Knutson et al., 2025; Lloyd-Jones et al., 2022; St-Onge et al., 2025). Suboptimal sleep patterns, such as short sleep duration, irregular (i.e., inconsistent) sleep timing, or poor sleep efficiency (i.e., fragmented sleep), exhibit well established associations with excess body mass, hypertension, and cardiovascular disease risk (Cappuccio et al., 2010; Li & Shang, 2021; Nagai et al., 2010; Tao et al., 2021; Yin et al., 2017). Professional sleep societies recommend adults obtain at least seven hours of sleep per night and maintain regular sleep schedules (timing and duration) to promote health and performance (Sletten et al., 2023; Watson et al., 2015). However, a substantial proportion of US adults report sleep patterns that fall short of meeting these recommendations (Lloyd-Jones et al., 2022; St-Onge et al., 2025). Several studies document the prevalence of self-reported short sleep duration (i.e., <6-7 hours per night) has increased over the last two decades (Khubchandani & Price, 2020; Sheehan et al., 2018; Wang et al., 2022), and recent data demonstrate ∼30% of adults report inadequate sleep or difficulties sleeping (Di et al., 2022; Pankowska et al., 2023). Additionally, sleep regularity, which represents the night-to-night variation in sleep timing or duration (Culver et al., 2025; Fischer et al., 2021), is an important dimension of sleep health that may be a stronger predictor of mortality risk than average sleep duration (Huang et al., 2025; Huang & Redline, 2019; Knutson et al., 2025; Makarem et al., 2020; Rogers et al., 2024; Sletten et al., 2023; Zhu et al., 2022) . For example, there are data demonstrating poor sleep regularity is a stronger predictor of coronary artery calcium (Full et al., 2023) and mortality risk (Windred et al., 2023) than short average sleep duration.

Existing literature suggests emerging adults (the transition from adolescence to adulthood; 18 - 29 years) (Scott et al., 2025) disproportionately demonstrate poorer health behaviors compared to other age groups in adulthood. For example, poorer sleep health is evidenced by observances of 40-70% of emerging adults reporting short sleep duration and high sleep irregularity (Dillon et al., 2015; Jonasdottir et al., 2021; Mbous et al., 2022; Pankowska et al., 2023; Park et al., 2019; Peltzer & Pengpid, 2016). Importantly, poor sleep may influence diet and physical activity, behaviors which can support overall health. Indeed, diet is a major modifiable risk factor for cardiometabolic health and chronic illness (Johnson, 2019). Poor diet quality is prevalent among emerging adults, with this age group consistently falling below recommended intakes for key nutrients in population-level data (Cowan-Pyle et al., 2024). Since establishing positive health behaviors early in the lifecourse has implications for long term cardiometabolic health, understanding sleep patterns in this population is a growing public health priority (Chaput et al., 2020; Itani et al., 2017; Miner & Kryger, 2017).

To our knowledge, there are limited data on multidimensional sleep and holistic diet quality. Studies on following a Mediterranean-style (Campanini et al., 2017; Godos et al., 2024; Verkaar et al., 2024) or plant-based diet (St-Onge et al., 2018) suggest a beneficial effect on sleep quality due to the rich composition of healthy fats, lean protein, fiber, and antioxidants. However, results vary based on population and measures used. Thus, there is a need for rigorous methodologies such as actigraphy and multi-day food and fluid diaries to assess diet. Therefore, we sought to examine associations between objective sleep (duration, efficiency, timing, regularity) and subjective sleep health with diet quality (DASH, HEI scores) in emerging adults. We hypothesized that individuals with worse sleep would report lower diet quality. This secondary, preliminary analysis will provide important foundational data linking multidimensional sleep health with habitual diet quality in emerging adults, offering insight that can be used to inform larger, more comprehensive studies and future interventions.

## Materials and Methods

### Study Design and Participants

Data were previously collected in four separate studies (total n = 164) and coalesced for these analyses (NCT04576338: n = 54, NCT04334135: n = 33, NCT04244604: n = 38, NCT05918744: n = 39). The procedures and protocols were approved by the Institutional Review Board at Auburn University and conducted in accordance with the Declaration of Helsinki. Data were collected by a single laboratory using the same data collection methods including sleep actigraphy devices, sleep questionnaires (e.g., Pittsburgh Sleep Quality Index; **PSQI**), and multi-day food and fluid diaries, which permitted pooling into a larger cohort. Each parent study had its own purpose and hypotheses separate from the current objective of evaluating associations between habitual sleep health and diet quality (Culver et al., 2025; Jeong et al., 2024; Linder et al., 2024; Robinson et al., 2023). The current research question was not part of our *a priori* analytic plan when we designed these studies. Thus, we acknowledge this study is a secondary data analysis. Nonetheless, all participants provided their verbal and written consent to undergo sleep and diet assessment. As we sought to examine sleep and diet among emerging adults, we excluded individuals younger than 18 and older than 29 from these analyses (Scott et al., 2025).

Participants were recruited from the Auburn-Opelika, Alabama area, primarily Auburn University. Participants provided a medical history during an initial screening visit. Participants self-reported their biological sex and gender, race, ethnicity, and the race and ethnicity of their parents (Culver et al., 2025). Participant’s body mass and height were recorded using a digital scale and stadiometer (Tanita WB-3000) and used to calculate body mass index (**BMI;** kg/m^2^). Individuals were excluded for the following reasons: history of any overt or uncontrolled chronic diseases including cardio/cerebrovascular, renal, metabolic, autoimmune, or malignancies; BMI ≥35; a recent history of COVID-19 infection; currently pregnant or breastfeeding; or diagnosis of a sleeping disorder (e.g., insomnia, obstructive sleep apnea, restless legs syndrome).

### Sleep Measures

#### Objective sleep

Participants were instructed to wear a Phillips ActiWatch Spectrum PLUS actigraph (Phillips Respironics, Bend, OR) for objective sleep monitoring on the dorsal side of the preferred wrist prior to coming to the laboratory for the experimental visit. The intended observation period ranged from 7-14 consecutive days across the four original studies. Participants with a minimum of six nights of observation were included in the present analysis. The actigraphy devices are validated against polysomnography, the gold standard assessment of sleep (e.g., duration; sensitivity >0.90, accuracy >0.80) (Kahawage et al., 2020; Quante et al., 2018). All objective sleep data were downloaded and analyzed in 60-second epochs. Rest intervals were coded through visual inspection of the accelerations in the daily actograms within the Actiware software along with the information from the event markers and the nightly sleep logs provided by the participants. Briefly, our team used a decision tree to determine sleep and wake times. Participants completed a sleep diary recalling the prior night’s sleep (bedtime, sleep onset latency, waketime, and time out of bed). The Philips Actiwatch Spectrum Plus also includes a self-reported event marker, and participants were trained in its use prior to leaving the lab. Because event markers are sometimes missed or recorded incorrectly, our procedure integrated sleep diary entries with actigraphy accelerometry patterns observed on the actogram to establish rest intervals. If the monitor was removed, participants were asked to record the date and time of removal on their sleep diary. Lastly, upon visual inspection, sleep data were manually adjusted with the consideration of the subjective information by an investigator (M.N. Culver) (Culver et al., 2025; Culver et al., 2022).

Sleep duration was defined as the amount of sleep obtained for each night (i.e., coincided with the longest sleep bout per 24 hours) the sleep watch was worn (Culver et al., 2022; St-Onge et al., 2025) and averaged to determine mean values (St-Onge et al., 2025). Sleep efficiency, a measure of sleep initiation and maintenance, was calculated using the following formula: (time asleep during longest rest period / duration of longest rest period) x 100 and averaged across nights (St-Onge et al., 2025). Sleep timing was comprised of three measures: bedtime, waketime, and midpoint of sleep. Bedtime was operationalized as the first epoch of sustained inactivity following the participant’s reported time they prepared for sleep, and waketime as the last epoch prior to sustained activity per the participant’s reported time they awoke, as determined by the actigraphy scoring algorithm and investigator confirmation. Sleep midpoint was defined as the clock time halfway between when an individual fell asleep and woke up. As described previously (Culver et al., 2025; Culver et al., 2022; Grosicki et al., 2023; Hoopes et al., 2021), sleep regularity was defined as the standard deviation (**SD**) of duration (i.e., duration SD), bedtime (i.e., bedtime SD), waketime (i.e., waketime SD), and midpoint (i.e., midpoint SD).

#### Subjective sleep quality

All participants completed the 19-item PSQI to characterize their sleep quality and disturbances over the previous month. The PSQI generates seven component scores (subjective sleep quality, latency, duration, efficiency, disturbances, medication use, and daytime dysfunction), which are summed to yield a global score of sleep quality. The PSQI is scored 0 to 21, where a value closer to zero is interpreted as greater self-reported sleep quality (Buysse et al., 1989; Culver et al., 2025).

#### Sleep adequacy

Sleep adequacy was determined by whether a participant met established definitions of multidimensional sleep health (St-Onge et al., 2025): duration between 6-8 hours as measured by actigraphy, efficiency ≥85%, timing (midpoint) between 2-4 am, and regularity (within-person SD of bedtime, waketime, and midpoint ≤ 60 minutes or 1 hour). Duration regularity adequacy was adapted for an emerging adult population and consistent with previously published literature, where adequate duration SD ≤1.2 hours (Hoopes et al., 2021). An optimal PSQI global score was ≤5 (St-Onge et al., 2025). As bedtime and waketime do not have established adequacy standards, they were excluded from this sub-analysis.

### Dietary Measures

#### Nutrient analysis

Participants were instructed to complete a minimum of three days of a food and fluid diary, consisting of at least two weekdays and one weekend day so the proportion of weekday:weekend days was the same for all studies. Two studies (NCT04334135 and NCT04244604) utilized two three-day food and fluid diaries, making a total of ≥six days of diet recording, while the other two (NCT04576338 and NCT05918744) utilized one three-day food and fluid diary. To help with estimating portion sizes, participants were provided verbal instructions pertaining to detail needed when reporting food and fluids (e.g., describing type and quantity of bread and meat, condiments, vegetables, cheese, etc. vs. simply reporting ham sandwich) along with a handout describing portion sizes, how to interpret nutrition labels, and sample food and fluid diaries as an example of the level of detail, similar to our prior studies using these approaches (Culver et al., 2025; Grosicki et al., 2023; Jeong et al., 2024; Robinson et al., 2023; Stute et al., 2025). The food and fluid diaries were returned at the laboratory testing visits and reviewed by a laboratory member with the participant present to obtain additional clarification. Diet records were analyzed for macro- and micronutrient intakes using the Nutrition Data System for Research (**NDSR**; University of Minnesota, Minneapolis, MN, USA) software. Analyses were conducted by a registered dietitian (S.O. Sanchez) or by trained staff under the dietitian’s supervision, with all entries checked by the dietitian. Data is reported as the average nutrient intake per 24-hour period from the food and fluid diaries.

#### Diet quality

Diet quality is frequently quantified using composite dietary quality indices, such as Dietary Approaches to Stop Hypertension (**DASH**) and the Healthy Eating Index (**HEI**) scores, both of which are aligned with the primary dietary guidance in the US, the Dietary Guidelines for Americans (**DGA**s) (Chiuve et al., 2012; *Dietary Guidelines for Americans, 2020-2025*, 2020; Miller et al., 2013; Reedy et al., 2014; Reedy et al., 2018; Shams-White et al., 2023; Soltani et al., 2020). The DASH pattern is recommended by the DGAs due to emphasizing fruits, vegetables and whole grains, low or fat free dairy, lean meat, legumes, while limiting added fat, sodium, and sugar, and has been studied as an effective diet pattern to reduce blood pressure and improve cardiometabolic health (*Dietary Guidelines for Americans, 2020-2025*, 2020; Phillips et al., 2019; Sacks et al., 2001; Siervo et al., 2015). For DASH adherence, we followed the DASH diet index developed by Mellen et al that focuses on nine target nutrients, of which five that should be higher (i.e., protein, fiber, magnesium, calcium, and potassium) and four that should be lower (i.e., total fat, saturated fat, sodium, and cholesterol) to determine greater adherence to the DASH diet (Fung et al., 2010; Liang et al., 2020; Mellen et al., 2008). Mellen et al selected sodium based on a recommendation of 2,400 mg, however we used 2,300 mg for our criteria as this is the sodium content recommended by the DGAs and was the target in the original DASH-Sodium diet trial per a 2,100 calorie diet (Svetkey et al., 1999). Micronutrients were indexed to per 1,000 calories. If a participant met the criteria for a given nutrient, the nutrient was assigned a value of one point. If they consumed a value halfway between the DASH diet target and control diet per the original DASH study, the nutrient was assigned a half point. For example, the DASH target for potassium was 2238mg/1000 Calories, the control diet target was 810 mg/1000 Calories, and the midpoint was 1524mg/1000 Calories. Because potassium is considered favorable, consuming above 1524 mg/1000 Calories would result in 0.5 points. In contrast, consuming below the midpoint would be considered favorable for a nutrient like sodium. If a participant’s intake fell on the unfavorable side of the midpoint (below it for nutrients that should be higher, or above it for nutrients that should be lower), the nutrient was assigned a zero. The total DASH diet score was generated by summing all nutrient components for a minimum of zero and a maximum of nine points, where a higher score indicated greater compliance with the DASH diet.

Separately, we reported HEI scores computed by NDSR from participants’ food and fluid diaries, as previously described (Reedy et al., 2018; Shams-White et al., 2023). Briefly, developed and validated by the National Cancer Institute and the US Department of Agriculture (Miller et al., 2011), the HEI is a measure of how well an individual’s intake aligns with the DGAs and is a proxy measure for diet quality. The HEI score includes 13 components: nine adequacy components result in higher scores for greater intake (total fruits, whole fruits, total vegetables, greens and beans, whole grains, dairy, total protein foods, seafood and plant proteins, fatty acids), and four moderation components result in higher scores for lower intake (refined grains, sodium, added sugars, saturated fats) (Shams-White et al., 2023). Scores are calculated on a density basis relative to energy intake, except for fatty acids which uses the ratio of polyunsaturated and monounsaturated to saturated fats. Component scores sum to a total of zero to 100, with higher values reflecting better overall diet quality. For this manuscript we focus on the total score as a global measure of diet quality. HEI scores were based on the most recently available HEI criteria, HEI-2020, which are meant to align with the 2020-2025 version of the DGAs (Shams-White et al., 2023).

### Statistical Analyses

We inspected all variables for normality using the Shapiro-Wilk test and QQ plots to compare the shapes of distributions. Descriptive characteristics are presented as means and standard deviations (mean ± SD) for normally distributed data or as medians and interquartile ranges (median [IQR]) for non-normally distributed data. The race variable was coded as White as the referent variable (i.e., coded as zero), Black as 1, and other races as 2. Biological sex was coded as male as the referent variable (i.e., coded as zero) and female as 1. We assessed diet scores among participants based on their sleep health (i.e., duration, efficiency, midpoint, duration SD, bedtime SD, waketime SD, midpoint SD, and PSQI). Our primary analysis entailed hierarchical regression models, as we’ve previously utilized (Culver et al., 2025; Tharpe et al., 2023), to examine associations between sleep health and diet quality. We utilized unadjusted linear regression models (model 1) and linear regression models that included sex, BMI, and race (model 2). We report unstandardized beta coefficients (B) and 95% confidence intervals [95% CI] for the regressions. Additionally, to compare groups, we compared participant DASH and HEI scores based on “adequate” and “inadequate” sleepers for each sleep dimension. We then utilized Mann-Whitney U tests to compare diet quality between sleep groups. Statistical significance for all analyses was defined *a priori* as *p* ≤ 0.05. Statistical analyses were performed using jamovi 2.3.28, RStudio 4.5.0 and figures were made with GraphPad Prism 10.6.1 (GraphPad, San Diego, CA).

## Results

See CONSORT figure for participant flow chart (**Figure 1**). The total number of participants enrolled was 164 and the analytic samples varied based on the available data collected from each participant (e.g., actigraphy analytic sample size = 106, PSQI analytic sample size = 122). Participant demographics are presented in **Table 1**. A majority of participants were female (Actigraphy: 54.7%; PSQI: 52.4%) and White (Actigraphy: 57.5%; PSQI: 53.3%). Participant descriptive sleep and diet data are presented in **Table 2**. Participants’ median wear time was 8 [6] nights and food and fluid diaries were collected across 3 [3] days. Participants’ median DASH score was 1.5 out of nine points and mean HEI score was 50.3 out of 100 points.

**Figure 1.**
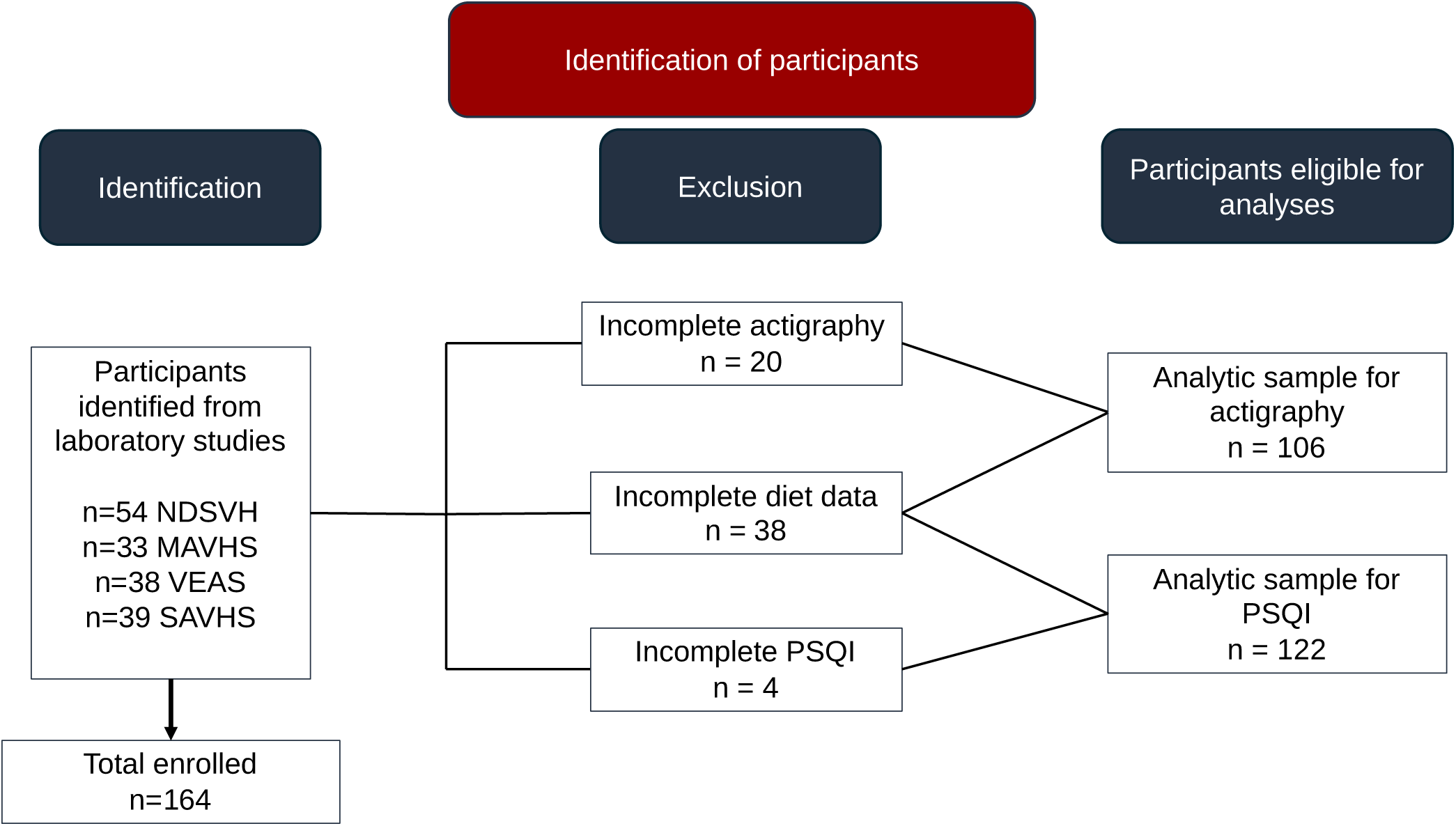
CONSORT diagram showing participant flow. Each study and corresponding ClinicalTrials.gov ID are as follows: “Neighborhood Disadvantage, Sleep and Vascular Health” (NDSVH); NCT04576338, “The Influence of Mitochondrial-Derived Reactive Oxygen Species on Racial Disparities in Neurovascular Function” (MAVHS); NCT04334135, “Vascular Effects of Acute Sodium” (VEAS); NCT04244604, and “Sleep and Vascular Health Study” (SAVHS); NCT05918744.

**Table 1.** Participant Demographics.

| **Table 1. Participant Demographics** | **Actigraphy**<br>**n=106** | **PSQI**<br>**n=122** |
| --- | --- | --- |
| Biological sex (Male; Female) | 48; 58 | 58; 64 |
| Age in years | 22, 2 | 22, 2 |
| Height, cm | 168.2, 15.5 | 169.5, 15.9 |
| Mass, kg | 72.4 ± 12.8 | 73.8 ± 13.9 |
| BMI, kg/m<sup>2</sup> | 24.2, 4.6 | 24.2, 5.1 |
| Race | n (%) | n (%) |
| White | 61 (57.5) | 65 (53.3) |
| Black | 32 (30.2) | 40 (32.8) |
| Other | 13 (12.3) | 17 (13.9) |
| Note: Data are presented as mean ± standard deviation for normally distributed data or median, IQR for non-normally distributed data. PSQI = Pittsburgh Sleep Quality Index. |  |  |

**Table 2.**
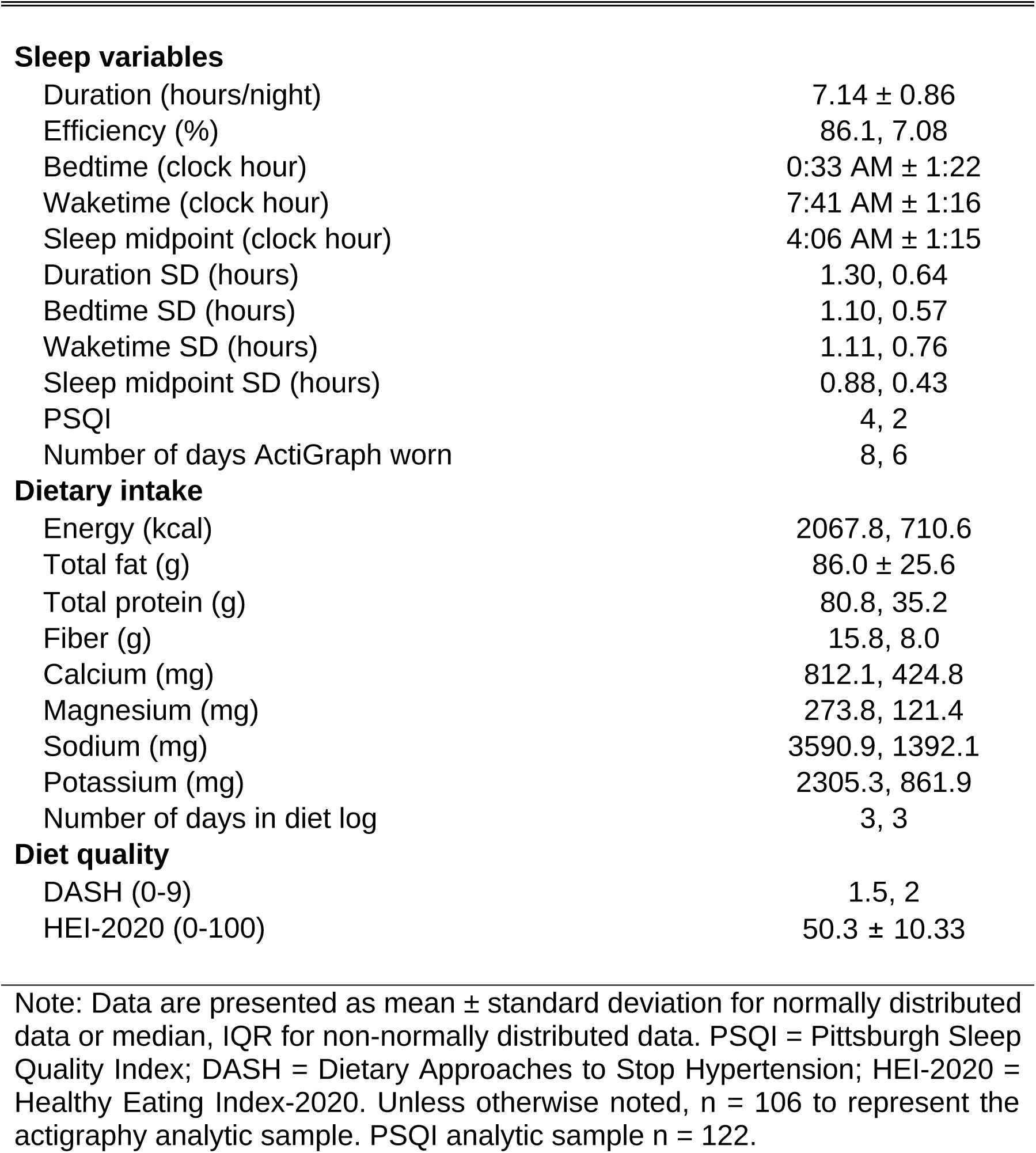
Participant sleep and self-reported dietary intake.

|  |  |
| --- | --- |
| Duration (hours/night) | 7.14 ± 0.86 |
| Efficiency (%) | 86.1, 7.08 |
| Bedtime (clock hour) | 0:33 AM ± 1:22 |
| Waketime (clock hour) | 7:41 AM ± 1:16 |
| Sleep midpoint (clock hour) | 4:06 AM ± 1:15 |
| Duration SD (hours) | 1.30, 0.64 |
| Bedtime SD (hours) | 1.10, 0.57 |
| Waketime SD (hours) | 1.11, 0.76 |
| Sleep midpoint SD (hours) | 0.88, 0.43 |
| PSQI | 4, 2 |
| Number of days ActiGraph worn | 8, 6 |
| **Dietary intake** |  |
| Energy (kcal) | 2067.8, 710.6 |
| Total fat (g) | 86.0 ± 25.6 |
| Total protein (g) | 80.8, 35.2 |
| Fiber (g) | 15.8, 8.0 |
| Calcium (mg) | 812.1, 424.8 |
| Magnesium (mg) | 273.8, 121.4 |
| Sodium (mg) | 3590.9, 1392.1 |
| Potassium (mg) | 2305.3, 861.9 |
| Number of days in diet log | 3, 3 |
| **Diet quality** |  |
| DASH (0-9) | 1.5, 2 |
| HEI-2020 (0-100) | 50.3 ± 10.33 |

Note: Data are presented as mean ± standard deviation for normally distributed data or median, IQR for non-normally distributed data. PSQI = Pittsburgh Sleep Quality Index; DASH = Dietary Approaches to Stop Hypertension; HEI-2020 = Healthy Eating Index-2020. Unless otherwise noted, n = 106 to represent the actigraphy analytic sample. PSQI analytic sample n = 122.

Unadjusted regression results (Models 1) and adjusted regression results (Models 2) are reported in **Tables 3-6**. Associations between sleep timing, sleep timing regularity measures, and PSQI with DASH scores are presented in **Tables 3**-**4**. When simple bivariate regressions were conducted for Model 1, earlier bedtime and earlier midpoint were associated with increased DASH (**Table 3**). Similar patterns were observed for greater efficiency, earlier waketime, and lower bedtime variability, though confidence intervals included zero. In adjusted models, earlier bedtime and earlier sleep midpoint (**Table 3)** retained their associations with higher DASH scores, and lower bedtime SD (**Table 4)** was also associated with higher DASH score..

**Table 3.** Sleep duration and timing predictors of DASH.

| Independent variables | Model 1<br>B(SE) | 95% CI | *p* | Model 2<br>B(SE) | *p* | 95% CI | *R*<sup>2</sup> | Overall<br>*p* |
| --- | --- | --- | --- | --- | --- | --- | --- | --- |
| Duration (hours) | 0.085 (0.127) | -0.167, 0.337 | 0.504 | 0.125 (0.117) | 0.288 | -0.107, 0.358 | 0.232 | <0.001 |
| Sex |  |  |  | 0.236 (0.199) | 0.240 | -0.160, 0.631 |  |  |
| BMI |  |  |  | -0.006 (0.029) | 0.832 | -0.064, 0.052 |  |  |
| Race (Black v White) |  |  |  | -0.410 (0.232) | 0.081 | -0.871, 0.051 |  |  |
| Race (Other v White) |  |  |  | 1.308 (0.316) | <0.001 | 0.681, 1.935 |  |  |
| Efficiency | 0.033 (0.017) | -0.001, 0.067 | 0.057 | 0.007 (0.017) | 0.680 | -0.026, 0.040 | 0.225 | <0.001 |
| Sex |  |  |  | 0.236 (0.203) | 0.248 | -0.167, 0.638 |  |  |
| BMI |  |  |  | -0.005 (0.029) | 0.858 | -0.063, 0.053 |  |  |
| Race (Black v White) |  |  |  | -0.437 (0.234) | 0.065 | -0.902, 0.028 |  |  |
| Race (Other v White) |  |  |  | 1.220 (0.316) | <0.001 | 0.593, 1.846 |  |  |
| **Bedtime (clock hour)** | **-0.175 (0.078)** | **-0.330, -0.020** | **0.027** | **-0.165 (0.073)** | **0.025** | **-0.309, -0.021** | **0.262** | **<0.001** |
| Sex |  |  |  | 0.191 (0.197) | 0.333 | -0.199, 0.582 |  |  |
| BMI |  |  |  | -0.002 (0.029) | 0.943 | -0.059, 0.055 |  |  |
| Race (Black v White) |  |  |  | -0.348 (0.229) | 0.132 | -0.802, 0.106 |  |  |
| Race (Other v White) |  |  |  | 1.333 (0.307) | <0.001 | 0.725, 1.942 |  |  |
| Waketime (clock hour) | -0.166 (0.086) | -0.336, 0.004 | 0.056 | -0.128 (0.079) | 0.107 | -0.284, 0.028 | 0.244 | <0.001 |
| Sex |  |  |  | 0.217 (0.199) | 0.276 | -0.177, 0.611 |  |  |
| BMI |  |  |  | -0.002 (0.029) | 0.955 | -0.059, 0.056 |  |  |
| Race (Black v White) |  |  |  | -0.418 (0.228) | 0.069 | -0.870, 0.033 |  |  |
| Race (Other v White) |  |  |  | 1.247 (0.308) | <0.001 | 0.637, 1.857 |  |  |
| **Midpoint (clock hour)** | **-0.189 (0.086)** | **-0.359, -0.018** | **0.031** | **-0.167 (0.079)** | **0.038** | **-0.325, -0.010** | **0.257** | **<0.001** |
| Sex |  |  |  | 0.193 (0.198) | 0.331 | -0.199, 0.585 |  |  |
| BMI |  |  |  | -0.000 (0.029) | 0.989 | -0.058, 0.057 |  |  |
| Race (Black v White) |  |  |  | -0.385 (0.227) | 0.093 | -0.836, 0.065 |  |  |
| Race (Other v White) |  |  |  | 1.295 (0.306) | <0.001 | 0.688, 1.902 |  |  |

Associations between sleep timing variables with the DASH score. Sleep measures were derived via actigraphy. Covariates: sex (where male is the referent), BMI, race (where White is the referent). Data presented include the diet variable regressed on its respective sleep predictor. SD = standard deviation, B = non-standardized beta, SE = standard error.

**Table 4.**
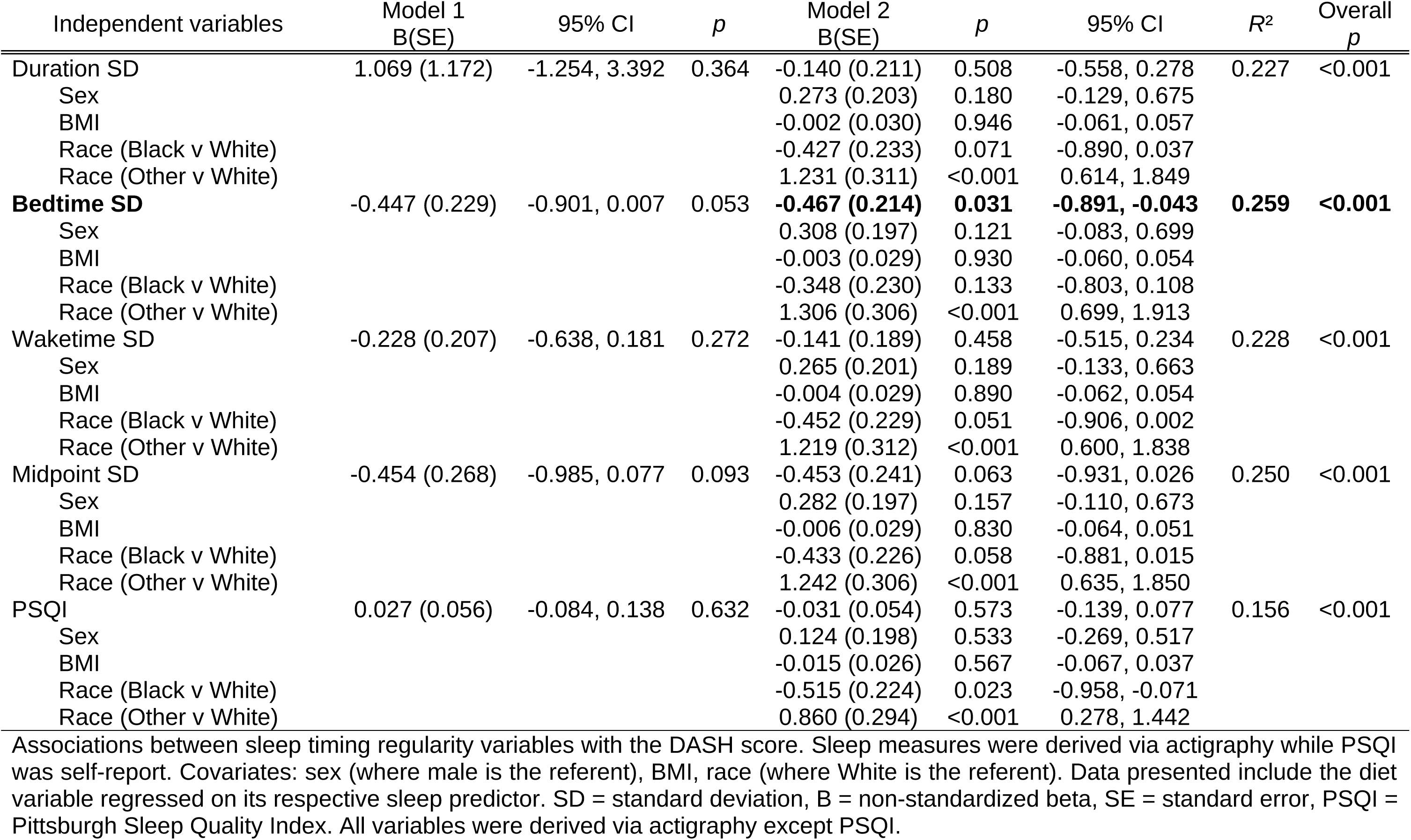
Sleep regularity and self-report quality predictors of DASH.

**Table 5.** Sleep duration and timing predictors of HEI.

| Independent variables | Model 1<br>B(SE) | 95% CI | *p* | Model 2<br>B(SE) | *p* | 95% CI | *R*<sup>2</sup> | Overall<br>*p* |
| --- | --- | --- | --- | --- | --- | --- | --- | --- |
| Duration (hours) | 1.069 (1.172) | -1.254, 3.392 | 0.364 | 1.296 (1.120) | 0.250 | -0.925, 3.517 | 0.180 | 0.001 |
| Sex |  |  |  | 2.930 (1.905) | 0.127 | -0.850, 6.710 |  |  |
| BMI |  |  |  | -0.340 (0.279) | 0.226 | -0.894, 0.214 |  |  |
| Race (Black v White) |  |  |  | -3.097 (2.220) | 0.166 | -7.500, 1.307 |  |  |
| Race (Other v White) |  |  |  | 8.970 (3.019) | 0.004 | 2.981, 14.959 |  |  |
| Efficiency | 0.274 (0.159) | -0.042, 0.590 | 0.088 | 0.060 (0.160) | 0.707 | -0.257, 0.377 | 0.171 | 0.002 |
| Sex |  |  |  | 2.957 (1.941) | 0.131 | -0.894, 6.807 |  |  |
| BMI |  |  |  | -0.330 (0.281) | 0.242 | -0.887, 0.227 |  |  |
| Race (Black v White) |  |  |  | -3.409 (2.241) | 0.131 | -7.855, 1.037 |  |  |
| Race (Other v White) |  |  |  | 8.096 (3.019) | 0.009 | 2.107, 14.086 |  |  |
| **Bedtime (clock hour)** | **-2.328 (0.705)** | **-3.726, -0.930** | **0.001** | **-2.180 (0.679)** | **0.002** | **-3.527, -0.833** | **0.247** | **<0.001** |
| Sex |  |  |  | 2.306 (1.838) | 0.212 | -1.340, 5.952 |  |  |
| BMI |  |  |  | -0.288 (0.268) | 0.285 | -0.819, 0.244 |  |  |
| Race (Black v White) |  |  |  | -2.144 (2.136) | 0.318 | -6.383, 2.094 |  |  |
| Race (Other v White) |  |  |  | 9.498 (2.862) | 0.001 | 3.819, 15.176 |  |  |
| **Waketime (clock hour)** | **-2.255 (0.776)** | **-3.794, -0.717** | **0.004** | **-1.871 (0.738)** | **0.013** | **-3.336, -0.407** | **0.220** | **<0.001** |
| Sex |  |  |  | 2.599 (1.864) | 0.166 | -1.100, 6.298 |  |  |
| BMI |  |  |  | -0.277 (0.273) | 0.313 | -0.819, 0.265 |  |  |
| Race (Black v White) |  |  |  | -3.021 (2.138) | 0.161 | -7.263, 1.221 |  |  |
| Race (Other v White) |  |  |  | 8.367 (2.888) | 0.005 | 2.636, 14.098 |  |  |
| **Midpoint (clock hour)** | **-2.616 (0.772)** | **-4.147, -1.086** | **<0.001** | **-2.347 (0.739)** | **0.002** | **-3.813, -0.881** | **0.246** | **<0.001** |
| Sex |  |  |  | 2.281 (1.841) | 0.218 | -1.371, 5.933 |  |  |
| BMI |  |  |  | -0.261 (0.269) | 0.333 | -0.794, 0.272 |  |  |
| Race (Black v White) |  |  |  | -2.582 (2.115) | 0.225 | -6.777, 1.613 |  |  |
| Race (Other v White) |  |  |  | 9.042 (2.850) | 0.002 | 3.387, 14.696 |  |  |

Associations between sleep timing variables with the HEI score. Sleep measures were derived via actigraphy. Covariates: sex (where male is the referent), BMI, race (where White is the referent). Data presented include the diet variable regressed on its respective sleep predictor. SD = standard deviation, B = non-standardized beta, SE = standard error.

**Table 6.** Sleep regularity and self-report quality predictors of HEI.

| Independent variables | Model 1<br>B(SE) | 95% CI | *p* | Model 2<br>B(SE) | *p* | 95% CI | *R*<sup>2</sup> | Overall<br>*p* |
| --- | --- | --- | --- | --- | --- | --- | --- | --- |
| Duration SD | -3.228 (2.007) | -7.208, 0.753 | 0.111 | -2.110 (2.009) | 0.296 | -6.097, 1.876 | 0.179 | 0.001 |
| Sex |  |  |  | 3.430 (1.931) | 0.079 | -0.402, 7.262 |  |  |
| BMI |  |  |  | -0.281 (0.284) | 0.324 | -0.843, 0.282 |  |  |
| Race (Black v White) |  |  |  | -3.127 (2.225) | 0.163 | -7.541, 1.286 |  |  |
| Race (Other v White) |  |  |  | 8.132 (2.967) | 0.007 | 2.246, 14.018 |  |  |
| Bedtime SD | -3.383 (2.128) | -7.602, 0.837 | 0.115 | -3.465 (2.062) | 0.096 | -7.556, 0.627 | 0.192 | <0.001 |
| Sex |  |  |  | 3.511 (1.904) | 0.068 | -0.266, 7.289 |  |  |
| BMI |  |  |  | -0.310 (0.277) | 0.266 | -0.861, 0.240 |  |  |
| Race (Black v White) |  |  |  | -2.773 (2.217) | 0.214 | -7.171, 1.624 |  |  |
| Race (Other v White) |  |  |  | 8.763 (2.952) | 0.004 | 2.906, 14.620 |  |  |
| Waketime SD | -3.703 (1.887) | -7.445, 0.038 | 0.052 | -3.108 (1.783) | 0.084 | -6.645, 0.428 | 0.194 | <0.001 |
| Sex |  |  |  | 3.408 (1.894) | 0.075 | -0.349, 7.166 |  |  |
| BMI |  |  |  | -0.302 (0.277) | 0.278 | -0.852, 0.248 |  |  |
| Race (Black v White) |  |  |  | -3.483 (2.162) | 0.110 | -7.772, 0.806 |  |  |
| Race (Other v White) |  |  |  | 7.791 (2.949) | 0.010 | 1.940, 13.641 |  |  |
| Midpoint SD | -4.289 (2.473) | -9.194, 0.616 | 0.086 | -4.421 (2.303) | 0.058 | -8.991, 0.149 | 0.199 | <0.001 |
| Sex |  |  |  | 3.389 (1.886) | 0.075 | -0.352, 7.131 |  |  |
| BMI |  |  |  | -0.340 (0.276) | 0.221 | -0.887, 0.208 |  |  |
| Race (Black v White) |  |  |  | -3.355 (2.158) | 0.123 | -7.635, 0.926 |  |  |
| Race (Other v White) |  |  |  | 8.293 (2.926) | 0.006 | 2.488, 14.099 |  |  |
| PSQI | -0.253 (0.505) | -1.253, 0.747 | 0.617 | -0.742 (0.498) | 0.139 | -1.728, 0.244 | 0.128 | 0.007 |
| Sex |  |  |  | 2.388 (1.815) | 0.191 | -1.207, 5.982 |  |  |
| BMI |  |  |  | -0.316 (0.242) | 0.194 | -0.794, 0.163 |  |  |
| Race (Black v White) |  |  |  | -4.001 (2.050) | 0.053 | -8.061, 0.060 |  |  |
| Race (Other v White) |  |  |  | 5.495 (2.687) | 0.043 | 0.173, 10.817 |  |  |

Associations between sleep timing regularity variables with the HEI score. Sleep measures were derived via actigraphy while PSQI was self-report. Covariates: sex (where male is the referent), BMI, race (where White is the referent). Data presented include the diet variable regressed on its respective sleep predictor. SD = standard deviation, B = non-standardized beta, SE = standard error, PSQI = Pittsburgh Sleep Quality Index. All variables were derived via actigraphy except PSQI.

Associations between sleep timing, sleep timing regularity measures, and PSQI with HEI scores are presented in **Tables 5-6**. When simple linear regressions were conducted for Model 1, earlier bedtime, waketime, and midpoint were all associated with higher HEI (**Table 5**), while lower waketime SD nearly met the criteria for statistical significance (**Table 6**). When covariates were included, the three sleep timing (i.e., later bedtime, waketime, midpoint) remained associated with HEI scores (**Table 5**). Sleep duration, efficiency, duration SD, bedtime SD, midpoint SD and PSQI held no associations with HEI in any of the models.

Participant sleep adequacy metrics are reported in **Figures 2-3**. In this analysis, 80.2% of participants from the overall sample demonstrated adequate sleep duration, 59.4% exhibited adequate efficiency, and 47.2% achieved an adequate sleep midpoint. Regarding sleep timing regularity, 39.6% of participants demonstrated adequate duration SD, 44.3% an optimal bedtime SD, 40.6% an adequate waketime SD, and 64.2% an adequate midpoint SD. Most participants reported adequate sleep, with 77.0% reporting ≤5 on the PSQI. Mann-Whitney U tests indicated higher DASH (**Figure 2h**) and HEI (**Figure 2i**) among participants obtaining an adequate compared with inadequate sleep midpoint. Additionally, DASH (**Figure 3h**) and HEI (**Figure 3i**) were higher among participants obtaining an adequate compared with an inadequate waketime SD.

**Figure 2.**
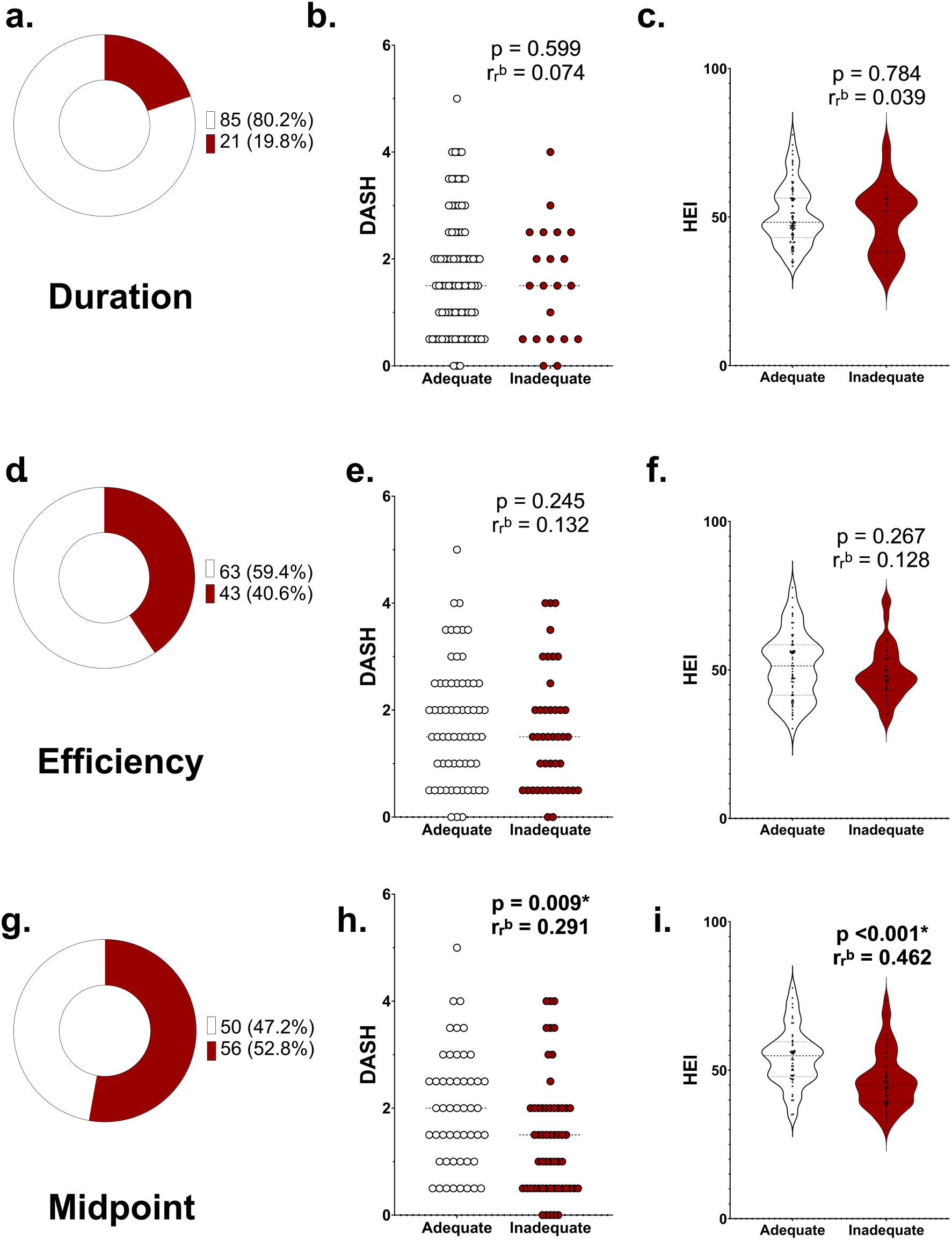
Participants’ diet data were split into optimal and non-optimal for each respective sleep metric with percentages reported in each donut graph. SD = standard deviation, DASH = Dietary Approaches to Stop Hypertension, HEI = Healthy Eating Index. Graphs 2a-c depict duration, 2d-f depict efficiency, 2g-i depict midpoint. Graphs 2a, 2d, 2g depict donut graphs from the overall sample with optimal in cream and non-optimal in crimson. Graphs 2b, 2e, and 2h represent Mann-Whitney U comparison tests for DASH scores. Graphs 2c, 2f, and 2i depict Mann-Whitney U comparison tests for HEI scores.

**Figure 3.**
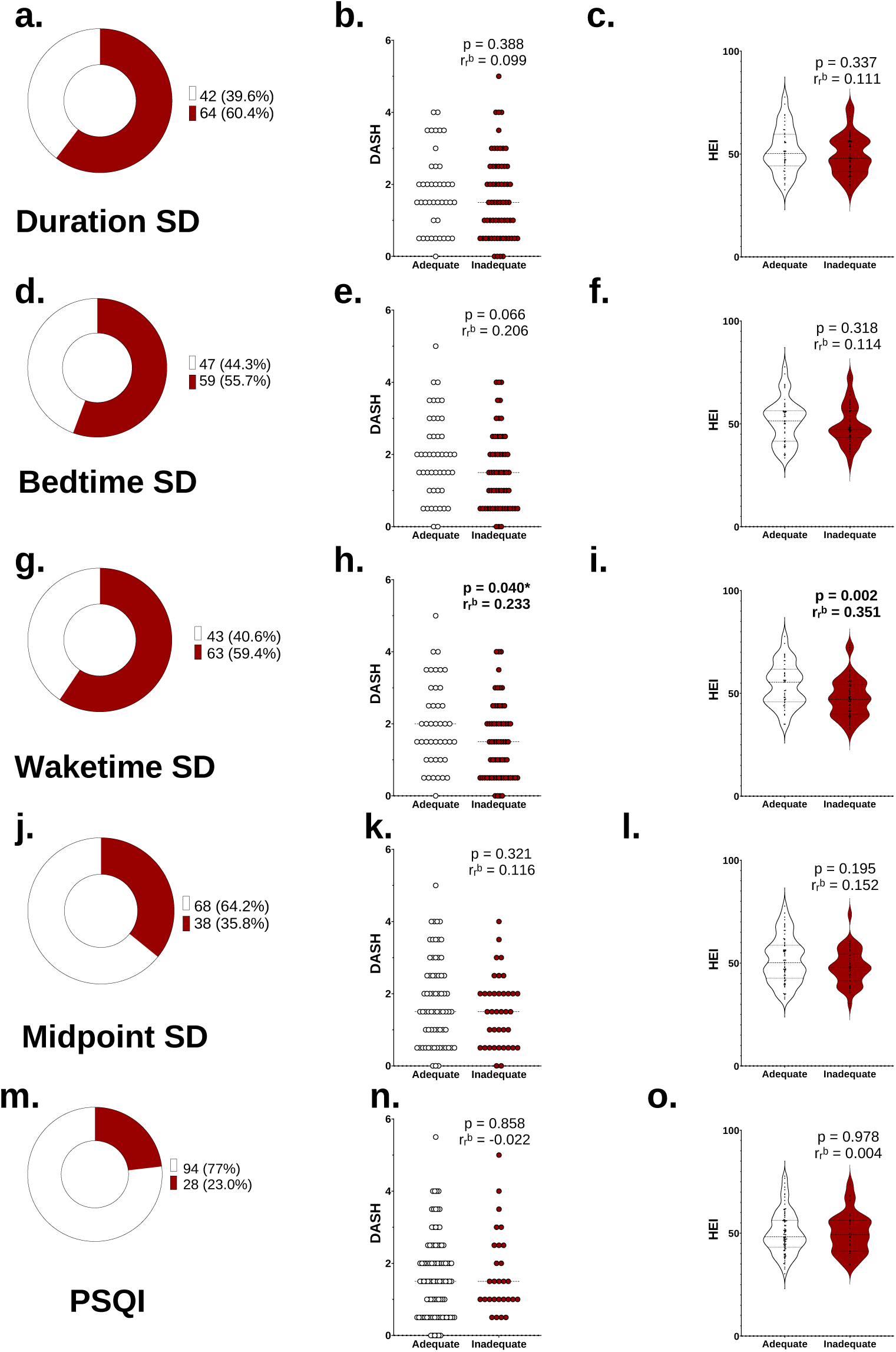
Participants’ diet data were split into optimal and non-optimal for each respective sleep metric with percentages reported in each donut graph. SD = standard deviation, DASH = Dietary Approaches to Stop Hypertension, HEI = Healthy Eating Index, PSQI = Pittsburgh Sleep Quality Index. Graphs 3a-c depict duration SD, 3d-f depict bedtime SD, 3g-i depict waketime SD, 3j-l depict midpoint SD, and 3m-o depict PSQI. Graphs 3a, 3d, 3g, 3j, 3m represent donut graphs from the overall sample with optimal in cream and non-optimal in crimson. Graphs 3b, 3e, 3h, 3k, and 3n represent Mann-Whitney U comparison tests for DASH scores. Graphs 3c, 3f, 3i, 3l, and 3o represent Mann-Whitney U comparison tests for HEI scores.

## Discussion

Our major findings demonstrate that in a sample of emerging adults, several measures of sleep health were associated with overall diet quality. Specifically, earlier bedtime, midpoint, and more consistent bedtime regularity were all independently associated with higher DASH scores. Similarly, we found earlier bedtime, waketime, midpoint, and more consistent midpoint regularity were all independently associated with higher HEI scores. Secondarily, we found when comparing adequate vs inadequate sleepers, individuals who achieved an optimal midpoint and waketime regularity exhibited better diet quality as measured by DASH and HEI. While we acknowledge the link between sleep health and diet quality is likely bidirectional (Knutson et al., 2025; St-Onge, Mikic, et al., 2016; St-Onge et al., 2012; St-Onge, Roberts, et al., 2016), these findings suggest that earlier and more regular sleep patterns are associated with better overall diet quality in emerging adults. Our findings advance previous literature by describing sleep and diet quality patterns, as well as further characterizing these relations among emerging adults (Zuraikat et al., 2021).

### Sleep duration

Our findings did not support an association between sleep duration and diet quality in emerging adults (e.g., **Table 3** and **Table 5**). These data are in contrast to several prior studies indicating specifically short sleep duration has an established influence on diet and is associated with diet quality and quantity (Patterson et al., 2014; Stern et al., 2014), even when controlling for body weight (Dashti et al., 2015). These published associations may be explained by shorter sleep meaning greater time spent awake and more opportunities for eating occasions. Suboptimal sleep duration may also promote hedonic seeking of energy dense foods and appetite dysregulation through hormones such as leptin, ghrelin, cortisol, and insulin (Godos et al., 2021). Regarding the discrepancy between our findings and prior studies, some of these studies used sleep restriction (e.g., <5.5 hours per night) in adults with habitually optimal sleep duration (Grandner, 2019) to demonstrate a role of duration. For example, in sleep restriction to four hours per night over two weeks, participants consumed an additional 300 calories ad libitum, presumably through snack foods (Covassin et al., 2022). Additionally, sleep restriction to 6.5 hours over two weeks led to increased consumption of higher glycemic index foods, with increases in dessert consumption, and one hour shorter sleep duration was associated with lower intake of fruit and vegetables the following day (Beebe et al., 2013; Winpenny et al., 2023). That experimental restriction studies artificially induce large within-person changes in sleep duration that exceed what is observed naturalistically in a cross-sectional design may explain the potential reasons for the lack of association in our sample. This further makes direct comparisons of effect sizes difficult.

However, these experimental findings of shorter sleep duration impacting diet are consistent with observational, population-level datasets. National Health and Nutrition Examination Survey (NHANES) data demonstrate associations between sleep duration and diet, where short sleep is associated with increased total caloric intake and intake from snack foods, lower food variety, and decreased intake of several micronutrients (e.g., vitamin C, lycopene) (Grandner et al., 2013; Kant & Graubard, 2014). Large population-based datasets like NHANES detect modest associations through greater sample heterogeneity and statistical power that may be difficult in a relatively homogenous (college students) and modest sample such as the current investigation.

### Sleep timing

Our regression models indicate that for every one hour earlier in bedtime, DASH increases by 0.17 points, for every one hour earlier in midpoint, DASH increases by 0.17 points (**Table 3**), and for every one hour earlier in bedtime SD, DASH increases by 0.47 points (**Table 4**). We also found that for every one hour earlier in bedtime, HEI increases by 2.18 points and for every one hour earlier in midpoint, HEI increases by 2.35 points. For every one hour earlier in waketime, HEI increases by 1.87 points (**Table 5**). There are mixed findings on the interplay among sleep timing, caloric intake, and diet quality (Hooker et al., 2020; Rusu et al., 2022). However, our sleep timing findings align well with published literature. Ideal sleep timing occurs when the dark period of the solar cycle (i.e., biologic night) aligns with one’s circadian clock (St-Onge et al., 2025), and an optimal definition has been established as the midpoint of sleep occurring between 2-4 am. Bedtime and sleep midpoint were closely interrelated in our cohort, reflecting that sleep midpoint is derived in part from bedtime. Accordingly these two variables produced similar associations with diet quality (e.g., a 0.17-point increase in DASH per one-hour-earlier shift in either).

In further support of optimal sleep midpoints, participants in our cohort who attained an adequate sleep midpoint demonstrated better DASH and HEI scores (**Figure 2**). The importance of sleep timing can be further illustrated by delayed sleep-wake phase disorder (DSWPD), or “night owl” syndrome, whereby one’s bedtimes occur between 2-6 am and waketimes are chronically delayed ≥ 2 hours from societal norms (Micic et al., 2026). The DSWPD is associated with excessive daytime sleepiness and reduced daytime functioning (Micic et al., 2026). Specific to diet, delayed sleep timing may promote hedonic regulation of appetite. For example, although individuals with DSWPD did not demonstrate any associations with diet quality in one study of adolescents (Berendsen et al., 2020), later sleep midpoints and skipping meals was associated with higher BMI in another study of adolescents (Lommi et al., 2024). These data also align with a study of Japanese women that reported those with the most delayed sleep midpoint, (corresponding to ∼5:31 am) were likely to engage in greater consumption of nutrients such as alcohol, fat, confections, and meat when compared to the earliest sleep midpoint group (2:32 am) (Sato-Mito et al., 2011).

### Sleep timing regularity

Our findings indicate greater sleep timing irregularity (e.g., bedtime SD and midpoint SD), but not duration irregularity, was associated with worse diet quality (**Tables 4 and 6**). In our study, sleep timing regularity variables were independently associated with diet quality. For every one hour increase in bedtime SD, DASH decreased by 0.47 points (**Table 4**). Additionally, for every one hour increase in midpoint SD, HEI decreased by 4.42 points (**Table 6**). These findings suggest more consistent sleep schedules are associated with better diet quality. This transitional life stage is characterized by lack of caregiver-directed structured support (i.e., parents/guardians, school schedules) while establishing independence, increased technology usage, or greater fluctuations in daily scheduling (e.g., socializing, studying) as possible explanations for poor sleep regularity (Brautsch et al., 2023; Lu et al., 2023; Park et al., 2006; Simon et al., 2025; Wood et al., 2018).

Individuals with an adequate waketime SD demonstrated better DASH and HEI (**Figure 3h-i**). Although a different population, these findings align with a study of adult women, where reducing bedtime irregularity over six weeks, even without extending sleep duration, improved adiposity and lowered leukocyte–platelet aggregates, suggesting that less sleep timing irregularity may support cardiometabolic health (St-Onge et al., 2020). Further recent research suggests sleep timing regularity may be more impactful for health than duration, where keeping consistent sleep-wake timing schedules were associated with a 20-48% lower risk of all-cause mortality in middle- and older-aged adults (Windred et al., 2023).

### PSQI

We did not observe any associations between PSQI and diet patterns. This finding is somewhat divergent from a previous study where participants who consumed higher calorie coffee drinks and energy drinks reported worse PSQI scores (Young et al., 2020), although this study used food frequency questionnaires as opposed to holistic diet. Other covariates were not predictive of sleep in any of the models we conducted except for race. Race is a complex social construct associated with sleep patterns, likely due to differential environmental exposures and psychosocial stress (Johnson et al., 2019). While more work is needed on racial and ethnic differences in sleep among emerging adults, in a study at an academic institution in the

Southeastern US, there were race-based differences in sleep measures, where White students reported the longest sleep duration, although greatest environmental sleep disturbances, when compared to other races (Yip et al., 2025). However, this study was conducted exclusively among students in their first semester of college.

### Holistic diet quality

The current investigation contributes to a growing body of literature seeking to understand holistic dietary patterns and their association with sleep as opposed to single nutrients (Campanini et al., 2017; Reid et al., 2019; Wu et al., 2019). Investigating sleep patterns in the context of a holistic dietary pattern such as DASH or HEI may provide a better framework with which to understand sleep health. Like diet, sleep is a modifiable health behavior that warrants further study among emerging adults (Scott et al., 2025).

Diet quality in our sample of emerging adults was low and aligned with prior literature. While there are limited data on emerging adult DASH diet adherence, the overall median DASH score in our sample was 1.5, lower than in a study of 56 young adults (age 26.7 ± 0.8 years) whose mean score was 2.7 (Lobene et al., 2022). An epidemiologic study found scores of 2.1 among healthy individuals younger than 45, and 2.8 among older ages (Park et al., 2017). One consideration is that our cohort was considerably younger than the individuals in these prior studies and the vast majority of the cohort was college students. Additionally, our sample’s HEI score was also aligned with that documented in other literature. Our mean HEI was 50.3, whereas other literature document it at 50.3 among emerging adults, and 56 among adults age 19-30 (Cowan-Pyle et al., 2024; *Dietary Guidelines for Americans, 2020-2025*, 2020). In our study, diet scores were higher among participants obtaining optimal and more consistent sleep. These data support the notion that public health recommendations to improve sleep may be a strategy to improve diet quality and overall cardiometabolic health, but additional data is needed to infer whether improving sleep health results in improved dietary habits.

This study is not without limitations. This was an observational analysis in a modest sample sized cohort. The sample was not large enough to adequately investigate diet among habitually short or long sleepers or race comparisons. Additionally, we cannot make causal inferences from this analysis. Building on our cross-sectional findings, future research should consider interventions targeting sleep timing regularity and its effect on diet quality and appetite regulation, which may assist in inferring causality. Nonetheless, the strengths of our investigation include the relatively comprehensive assessment of diet with multi-day food and fluid diaries, and our inclusion of self-reported and objective sleep measures to assess multidimensional sleep health in emerging adults.

## Conclusion

Our major findings demonstrate that in a sample of emerging adults, earlier bedtime, midpoint, and more consistent bedtime regularity were all independently associated with higher holistic diet scores. While we acknowledge the link between sleep health and diet quality is likely bidirectional, our data support the notion that earlier and more regular sleep patterns are associated with better overall diet quality in emerging adults.

## Data Availability

All data produced in the present study are available upon reasonable request to the authors

## Acknowledgements

SOS, MNC, and ATR designed research question; MNC and BAL conducted primary data collection; MNC and SOS performed sleep and nutrition data analysis. SOS conducted statistical analysis. SOS and ATR led writing of the first draft of the manuscript with subsequent edits from all authors. SOS had primary responsibility for final content. All authors engaged in data interpretation and read and approved the final manuscript.

## Declaration of generative AI use

The authors report generative AI was not used in the research or preparation of this manuscript.

